# Reliability and validity of the UK Biobank cognitive tests

**DOI:** 10.1101/19002204

**Authors:** Chloe Fawns-Ritchie, Ian J Deary

## Abstract

UK Biobank is a health resource with data from over 500,000 adults. The participants have been assessed on cognitive function since baseline. The cognitive tests in UK Biobank are brief and bespoke, and are administered without supervision on a touchscreen computer. Psychometric information on the tests is limited. The present study examined their concurrent validity and short-term test-retest reliability. A sample of 160 participants (mean age=62.59, SD=10.24) completed the UK Biobank cognitive assessment and a range of well-validated cognitive tests (‘reference tests’). Fifty-two participants returned 4 weeks later to repeat the UK Biobank tests. Correlations were calculated between UK Biobank tests and the reference tests. Four-week test-retest correlations were calculated for UK Biobank tests. UK Biobank cognitive tests showed a range of correlations with their respective reference tests, i.e. those tests that are thought to assess the same underlying cognitive ability (mean Pearson *r*=0.53, range=0.22 to 0.83, *p*≤.005). Four-week test-retest reliability of the UK Biobank tests were moderate-to-high (mean Pearson *r*=0.55, range=0.40 to 0.89, p≤.003). Despite the brief, non-standard nature of the UK Biobank cognitive tests, some showed substantial concurrent validity and test-retest reliability. These psychometric results provide currently-lacking information on the validity of the UK Biobank cognitive tests.

## Introduction

UK Biobank is a large prospective cohort study that was designed to investigate the health of middle-aged and older adults residing in the UK (https://www.ukbiobank.ac.uk/)^1^. At baseline (2006-2010), over half a million participants aged 40 to 70 years attended a UK Biobank clinic and completed a touchscreen questionnaire which collected information on early life factors, family history of disease, lifestyle, and health. A cognitive assessment was administered as part of this fully-automated touchscreen questionnaire. Physical measurements were taken, and blood, saliva and urine samples were collected in order to genotype participants and to assess biomarkers of disease^1^. Most participants also agreed to have their data linked to health records. Subsamples of UK Biobank participants have undergone repeat testing. One of the repeat studies involved 20,000 participants returning between 2009 and 2013 to complete the baseline assessment again. Currently, UK Biobank are conducting an imaging study (started in 2014) in which 100,000 participants are being asked to complete the baseline assessment again and undergo brain and body scanning. In addition to these clinic-based studies, subsamples have completed web-based assessments, including web-based cognitive testing. More information on the data collected at each assessment is reported elsewhere^1^.

The very large sample size and the breadth of information collected in UK Biobank, and the fact these data are available to researchers worldwide, makes this dataset a valuable resource for carrying out research into health and disease in ageing. At the time of writing, 1,463 research groups have applied to use the UK Biobank data (http://www.ukbiobank.ac.uk/approved-research/), and 762 research reports have been published using these data (https://www.ukbiobank.ac.uk/published-papers/).

Cognitive function has been assessed in UK Biobank since baseline. The original UK Biobank cognitive assessment was very brief (approximately 5 minutes). At baseline, almost all participants completed the Pairs Memory test, a test of visual memory, and the Reaction Time test, a measure of processing speed. Subsamples also completed tests of working memory (Numeric Memory test), prospective memory (Prospective Memory), and verbal and numerical reasoning (Fluid Intelligence)^1^. This test battery, excluding Numeric Memory, was then administered again at the repeat study that tested 20,000 people. Subsequently, more detailed measures of cognitive function assessing cognitive domains known to decline with increasing age^2^ were introduced at the imaging study. The new tests included tests of verbal declarative memory, executive function, and non-verbal reasoning. To estimate prior cognitive functioning, a measure of vocabulary was also included, which tends to remain relatively stable across the age range assessed in UK Biobank^2^. More information on the cognitive tests and domains assessed at each UK Biobank assessment are reported in Supplementary Table S1. The cognitive data, alongside the health, lifestyle, biomarker, genetic, and body and brain imaging data available in UK Biobank, provide materials for investigating the predictors and consequences of normal and pathological cognitive decline.

Cognitive functions are typically assessed under standardised conditions by a trained psychological tester, which may be seen as the gold standard method of cognitive assessment. Probably owing to the size of the UK Biobank sample, and the magnitude of other data being collected, the method of cognitive data collection was made efficient via a fully-automated touchscreen assessment. Moreover, the cognitive tests used in UK Biobank were brief measures, and were administered unsupervised^1^. Some of the UK Biobank cognitive tests were specifically designed for use in UK Biobank, whereas others were adapted versions of commonly-used tests which have been modified for use at the fully-automated touchscreen assessment.

Before the UK Biobank cognitive tests can be used with confidence to investigate healthy and pathological age-related cognitive changes, the psychometric properties of these tests should be investigated in more detail. One study^3^ examined the stability (test-retest reliability) of original UK Biobank tests over a four year interval between the baseline study and repeat study. It found that Fluid Intelligence (correlation between baseline and repeat *r* = 0.65) and Reaction Time (*r =* 0.54) showed substantial stability^3^. However, the test-retest reliability of Pairs Matching was low (*r* = 0.19)^3^.

The first aim of the present study was to examine the concurrent validity of the enhanced UK Biobank cognitive tests that are being administered to UK Biobank participants at the imaging study. An independent sample of participants who had not taken part in UK Biobank was recruited. They were administered the enhanced UK Biobank cognitive assessment. In addition, they were administered a battery of well-validated, standard cognitive tests; hereinafter we will call these ‘reference tests’. The concurrent validity of the UK Biobank tests was investigated by correlating scores on the UK Biobank tests with scores on the reference tests. For each of the UK Biobank tests, we chose a reference test that we judged was assessing the same underlying cognitive domain. We predicted that the correlations between UK Biobank tests and reference tests that assessed a similar cognitive domain should show higher correlations than those between UK Biobank tests and reference tests that assessed different cognitive domains.

The second aim of the present study investigated whether a component of general cognitive ability (*g*) was present in the correlations among the unsupervised UK Biobank tests, and whether any such *g* component correlated highly with a measure of general cognitive ability created using the reference tests administered by a trained tester under standardised conditions. One of the most replicated findings in psychological research is that performance on tests of cognitive function are positively correlated^4-7^. Principal component analysis (PCA) is often used to create a general measure of cognitive ability by combining scores on a wide range of cognitive tests into one composite general ability score. This composite score, which is created by saving scores based on the first unrotated principal component, typically accounts for about 40% of the variance in a wide range of different cognitive tests^8,9^. Lyall et al.^3^ investigated whether a general cognitive ability component could be extracted from the UK Biobank cognitive tests administered at baseline. In a PCA model including Reaction Time, Pairs Matching, Fluid Intelligence, and Numeric Memory, the first unrotated principal component accounted for 40% of the variance and the individual test scores all loaded at ≥ 0.49 on this component^3^. The enhanced UK Biobank cognitive assessment includes all baseline tests as well as more detailed tests introduced at the web-based and imaging studies (Supplementary Table S1). The present study expanded the work carried out by Lyall et al.^3^ and examined the covariance structure of the enhanced UK Biobank cognitive tests. Measures of general cognitive ability, created using completely different sets of tests of cognitive function have been found to correlate highly (*r* ≥ 0.77) in one study^6^ and almost perfectly (*r* ≥ 0.99) in another^5^. The present study predicted that the correlation between a *g* component created using the UK Biobank tests and a *g* component created using the reference tests would be high (e.g., *r* > 0.7).

The third aim of this study was to characterise the test-retest stability of all UK Biobank cognitive tests. Scores on reliable cognitive tests should be relatively stable over a short interval. Typical test-retest intervals are usually between 2-4 weeks. Lyall et al.^3^ tested the stability of the original UK Biobank tests over a 4 year interval. The test-retest interval in Lyall et al.^3^ was much longer than typical test-retest intervals, and could have included ageing effects, and, therefore, this might not be a useful indicator of the shorter-term stability of these tests. No one has examined the test-retest reliability of the newer UK Biobank tests.

## Methods

### Participants

Participants were identified through the University of Edinburgh Volunteer Panel and Join Dementia Research (https://www.joindementiaresearch.nihr.ac.uk/home?login). Both are databases of volunteers who are interested in taking part in research. Potential participants who were aged 40 to 80 years old—similar to the age range used in UK Biobank—and who were able to travel to the Psychology Department at the University of Edinburgh, were contacted and invited to take part in this study. UK Biobank participants and people with a diagnosis of dementia or mild cognitive impairment were not eligible for this study. A total of 160 participants were recruited. Informed content was obtained for all participants. This study received ethical approval from the University of Edinburgh Psychology Research Ethics Committee (reference number 2-1718/3). The methods reported in this study are in accordance with relevant guidelines and regulations.

### Materials

#### UK Biobank cognitive test battery

UK Biobank provided a stand-alone version of the UK Biobank cognitive test battery that was administered at the UK Biobank imaging study. To make the testing session as similar as possible to the UK Biobank clinic study sessions, the present study used the same touch screen monitor and computer setup as that which is used at the UK Biobank clinics’ imaging sessions. The UK Biobank cognitive assessment was designed to be fully-automated and the tests were administered unsupervised. There are UK Biobank staff in the clinic but participants are expected to sit and work through the assessment independently. Therefore, in the present study, participants were given brief oral instructions that they were going to complete some tasks on the computer on their own, and that they were to follow the instructions on the screen. One author (CF-R) was present in the room while participants completed the UK Biobank tests, but participants were left to work through the tests on their own. The tests administered as part of the UK Biobank cognitive assessment are listed in Table 1 and a detailed description of each test’s contents, administration, and scoring is provided in the Supplementary Methods.

**Table 1.** UK Biobank cognitive tests, general cognitive tests, and reference cognitive tests administered in the current study

| Cognitive domain | UK Biobank tests |  | General and reference tests |  |
| --- | --- | --- | --- | --- |
|  | Test name | Abbreviation | Reference test and source | Abbreviation |
| Visual declarative memory | Pairs Matching Test | UKB Pairs Matching | Wechsler Memory Scale IV Designs I and Designs II<sup>25</sup> | WMS-IV Designs I<br>WMS-IV Designs II |
| Processing speed | Reaction Time Test | UKB RT | Deary-Liewald Reaction Time Test Simple Reaction Time and Choice Reaction Time<sup>19</sup> | DLRT Simple RT<br>DLRT Choice RT |
| Prospective memory | Prospective Memory Test | UKB Prospective Memory | Rivermead Behavioural Memory Test - Extended Version Appointments<sup>26</sup> | RMBM Appointments |
| Verbal and numerical reasoning | Fluid Intelligence Test | UKB Fluid IQ | - | - |
| Working Memory | Numeric Memory Test | UKB Numeric Memory | Wechsler Adult Intelligence Scale IV Digit Span Forwards, Backwards and Sequence<sup>27</sup> | WAIS-IV Digit Span Forwards<br>WAIS-IV Digit Span Backwards<br>WAIS-IV Digit Span Sequence |
| Executive function | Trail Making Test parts A and B<sup>a</sup> | UKB TMT part A<br>UKB TMT part B | Trail Making Test parts A and B<sup>18</sup> | TMT part A<br>TMT part B |
| Processing speed | Symbol Digit Substitution Test | UKB Symbol Digit | Symbol Digit Modalities Test<sup>12</sup> | SDMT |
| Crystallised ability | Picture Vocabulary<sup>b</sup> | UKB Picture Vocabulary | Peabody Picture Vocabulary Test, Fourth Edition<sup>22</sup><br>National Adult Reading Test<sup>28</sup><br>NIH Toolbox Picture Vocabulary Test<sup>15</sup> | PPVT<br>NART<br>NIH Toolbox Picture Vocabulary |
| Verbal declarative memory | Paired Associate Learning Test<sup>c</sup> | UKB PAL | Wechsler Memory Scale IV Verbal Paired Associates I and Verbal Paired Associates II<sup>25</sup> | WMS-IV VPA I<br>WMS-IV VPA II |
| Executive function | Tower Rearranging Test<sup>d</sup> | UKB Tower Test | Delis-Kaplan Executive Function System Tower Test<sup>29</sup> | D-KEFS Tower Test |
| Non-verbal reasoning | Matrix Pattern Completion<sup>e</sup> | UKB Matrices | COGNITO Matrices<sup>24</sup> | COGNITO Matrices |
| Subjective memory | - | - | Self-rated memory questions<sup>30</sup> | Self-rated memory |
| Global cognitive function | - | - | Mini Addenbrooke's Cognitive Examination<sup>21</sup> | M-ACE |

<sup>a</sup>Adapted version of the Halstead-Reitan Trail Making Test<sup>18</sup><sup>b</sup>Adapted version of the NIH Toolbox Picture Vocabulary Test<sup>15</sup><sup>c</sup>Adapted version of the Test the Nation Paired Associate Learning Test<sup>31</sup><sup>d</sup>Adapted version of the one-touch Tower of London Test<sup>32</sup><sup>e</sup>Adapted version of the COGNITO Matrices test<sup>24</sup>

#### General tests

Brief screening tests of cognitive impairment, such as the Addenbrooke’s Cognitive Examination III (ACE-III)^10^, and measures of subjective memory complaints are often applied in studies of normal and pathological ageing. A cognitive screening test and a subjective memory questionnaire (see Table 1) were included in the current study to investigate the correlations between the UK Biobank cognitive tests and frequently-used measures of global cognitive function. Detailed descriptions of these tests are provided in the Supplementary Methods.

#### Reference tests

To be able to test the concurrent validity of each UK Biobank test, a battery of standard neuropsychological tests was administered. For each UK Biobank test, we selected one or more well-validated, standard cognitive test that resembles the UK Biobank test in terms of the underlying cognitive domain thought to be assessed, and in the actual content of the task (‘reference tests’). The reference tests were all administered under standardised conditions, one-to-one, face-to-face, by a trained tester, strictly following the administration instructions. We note the degree of similarity between each of the UK Biobank tests and the chosen reference tests varies. Whereas some of the reference tests use the same items as the UK Biobank tests, others are different versions of the same test, and others still are different tests that are thought to assess the same underlying cognitive ability. To be clear, some reference tests are better ‘matches’ for the UK Biobank tests than others, which should temper expectations about the respective UK Biobank-reference tests’ associations. The reference tests chosen for each UK Biobank test are shown in Table 1. A detailed description of each of the reference tests’ contents, administration, and scoring is provided in the Supplementary Methods. No reference test was chosen for the UKB Fluid IQ test because no test was identified by the authors as a suitable comparator.

#### Demographic and health questionnaire

Information on age, sex, and education was collected. Participant’s age was calculated from their date of birth. To measure education, participants were asked, “How many years of full-time education have you completed?”. General health was assessed by asking participants, “In general, would you say your health is excellent, very good, good, fair, or poor”, and “Compared to one year ago, how you would rate your health in general now?”. Participants selected from the following answers: Much better now than one year ago; somewhat better now than one year ago; about the same; somewhat worse now than one year ago; much worse now than one year ago.

#### UK Biobank cognitive assessment questionnaire

The UK Biobank cognitive test battery was designed to be administered unsupervised, and, although there were staff members working in the UK Biobank clinic during test administration, participants were expected to work through the cognitive assessment independently without a tester observing. Because no tester was there to help participants understand the test instructions, the onscreen instructions for each test must be clear. To assess this, after completing the UK Biobank cognitive assessment participants were asked, “Did you generally find that the instructions for all the tasks were clear?”. Participants answered either yes or no. Next, participants were shown screenshots of each UK Biobank cognitive test and were asked “Were the instructions for this test clear?”. Participants answered either yes or no.

The UKB Numeric Memory task was designed to assess backward digit span, which involves participants remembering a sequence of digits and then mentally reversing them in their mind. However, for this task, all the numbers in the to-be-remembered sequence were presented on the screen at once. This meant that some individuals were able to get the correct answer by reading the number sequence from right-to-left and not reversing the digits in their mind. These individuals are actually performing a forward digit span, which is an easier task. To identify the number of participants who completed this task forwards or backwards, participants were asked how they completed this task. The possible options were: read from left-to-right and reversed the digits in your mind; read from right-to-left and did not need to reverse digits in your mind; a mixture of both; something different.

### Procedure

The study visit for each participant in the present study took place in the Psychology Department at the University of Edinburgh. All assessments were administered by the same psychology-graduate tester (CF-R) in a quiet room, one-to-one, free of distractions. After reading the information sheet and signing the consent form, participants were administered the demographic and health questionnaire, and then the self-rated memory questionnaire. The testing session took approximately 2.5 to 3 hours to complete. The test order was counter-balanced. Individuals with even participant ID numbers completed the UK Biobank tests before completing the M-ACE and reference tests. Individuals with odd participant ID numbers completed the M-ACE and reference tests first and then completed the UK Biobank tests. The UK Biobank questionnaire was administered immediately after completing the UK Biobank cognitive assessment. Approximately half-way through the session, participants were given a short break. The test order for participants with odd and even ID numbers is shown in Supplementary Table S2.

Participants who indicated on the consent form that they would be willing to return for a second study visit, and who were able to arrange an appointment four weeks (± 1 week) after the first assessment, returned and completed the UK Biobank cognitive assessment again. Individuals who agreed to return for a second visit were administered the UK Biobank questionnaire after completing the UK Biobank tests for a second time.

### Statistical analyses

To examine the association between UK Biobank cognitive tests, general tests, and reference tests with basic demographic characteristics, correlations were calculated between all cognitive tests and age, sex, years of education, and self-reported general health. For all correlational analyses reported in this paper, both Pearson *r* and Spearman *rho* correlations were calculated. Point-biserial correlations were calculated for correlations with sex and UKB Prospective Memory, as these are binary variables. Concurrent validities of the UK Biobank tests were calculated by correlating the UK Biobank cognitive tests with the general test and reference tests. Partial correlations, adjusting for age, were also calculated to determine whether the sizes of the associations between UK Biobank cognitive tests and the general and reference tests remained after controlling for age. The correlation between scores on WMS-IV Designs I and II was high (*r* = 0.65, *p* < .001); therefore, a total score was created by summing the scores on Designs I and Designs II (WMS-IV Designs Total; max score = 240). Similarly, the correlation between WMS-IV VPA I and II was high (*r* = 0.89, *p* < .001); therefore, a total VPA score was created by summing the scores on VPA I and VPA II (WMS-IV VPA Total; maximum score = 70).

To investigate whether a measure of general cognitive ability created using UK Biobank cognitive tests was highly correlated with a measure of general cognitive ability created using some of the well-validated reference tests, three measures of general cognitive ability were created using the following combinations of tests: 1) all UK Biobank tests included in the enhanced assessment administered at the imaging study; 2) the UK Biobank baseline tests; and 3) a selection of the reference tests. These measures of general cognitive ability were created by entering cognitive tests scores into a PCA, checking the eigenvalues and scree plots, and saving the scores on the first unrotated principal component. Before cognitive test scores were entered into the PCA, test distributions (Supplementary Fig. S1) were inspected and, where possible, scores with non-normal distributions were transformed. The specific transformations performed are described below. To reduce the influence of any outliers, tests scores were winsorized to 3 SD.

#### General cognitive ability – using 11 reference tests

Scores on the following reference tests were entered into a PCA: TMT part B (log-transformed), SDMT, WMS-IV Designs Total, WAIS-IV Digit Span Total (created by summing scores on WAIS-IV Digit Span Forward and Digit Span Backward), D-KEFS Tower Test, DLRT Choice RT (log-transformed), NIH Toolbox Picture Vocabulary, NART (scores were reverted and log-transformed), PPVT (scores were reverted and log-transformed), WMS-IV VPA Total, and COGNITO Matrices score. Eigenvalues and scree plot (Supplementary Fig. S2) indicated two components. These two components accounted for 56% of the variance in the 11 reference cognitive tests. Test loadings (unrotated and rotated using oblique rotation) are shown in Supplementary Table S3. Scores on the first unrotated principal component, which accounted for 35% of the total variance, were saved and used as a measure of general cognitive ability (*g:reference-11*).

#### General cognitive ability – using 11 UK Biobank cognitive tests

Scores on the following tests were entered into a PCA: UKB Pairs Memory (log (x+1) transformed), UKB RT (log-transformed), UKB Prospective Memory, UKB Fluid IQ, UKB Numeric Memory, UKB TMT part B (log transformed), UKB Symbol Digit, UKB Picture Vocabulary, UK Paired Associate Learning, UKB Tower Test, and UKB Matrices. Eigenvalues and scree plot (Supplementary Fig. S3) indicated two components. These two components accounted for 46% of the variance in the 11 UK Biobank cognitive tests. Test loadings (unrotated and rotated using oblique rotation) are shown in Supplementary Table S4. Scores on the first unrotated principal component, which accounted for 34% of the total variance, were saved and used as a measure of general cognitive ability (*g:UKB-11*).

#### General cognitive ability – using 5 UK Biobank cognitive tests

Scores on the following tests were entered into a PCA: UKB Pairs Memory (log (x+1) transformed), UKB RT (log-transformed), UKB Prospective Memory, UKB Fluid IQ, and UKB Numeric Memory. Eigenvalues and scree plot (Supplementary Fig. S4) indicated one component. This component accounted for 38% of the total variance in the 5 tests. The test loadings are reported in Supplementary Table S5. Scores on this unrotated principal component were saved and used as a measure of general cognitive ability *(g:UKB-5).*

Correlations between the three measures of general cognitive ability were calculated. The correlations and age-adjusted correlations between *g:reference-11* and each of the UK Biobank cognitive tests were calculated. We also calculated the correlations and age-adjusted correlations between *g:UKB-11* and *g:UKB-5* with each of the general and reference tests.

To investigate whether participants thought the instructions for the UK Biobank cognitive tests were clear, the number and percentage of participants who answered ‘no’ to “Did you generally find that the instructions for all the tasks were clear?” was calculated. Next, the number and percentage of participants who reported ‘no’ when asked whether the instructions for each individual UK Biobank test were clear was calculated.

The number and percentage of participants who reported carrying out a forward digit span, a backward digit span, a mixture of both, or something else when completing UKB Numeric Memory was calculated. Between-group analysis of variance (ANOVA) was used to determine whether mean performance on the UKB Numeric Memory test differed by technique reported.

To measure the short-term stability of the UK Biobank tests, Pearson and Spearman test-retest correlations were calculated between scores on the UK Biobank tests at Time 1 and Time 2.

## Results

Participant characteristics are reported in Supplementary Table S6. A total of 160 participants (mean age = 62.59, SD = 10.24) completed the full assessment at Time 1. Of these, 52 participants (mean age = 61.69, SD = 9.70) returned and repeated the UK Biobank tests at Time 2. The mean time to repeat was 28.88 days (SD = 2.02, range = 26 to 36). The sample used here were relatively highly educated (mean years of full-time education = 16.19, SD = 2.73), and most reported their health to be very good (n = 85; 53.1%) or excellent (n = 36, 22.5%). Extended descriptive statistics (n, mean, SD and range) for each of the cognitive tests administered in this study are reported in Supplementary Table S7.

For all correlations reported throughout this report, both Pearson correlations and Spearman rank-order correlations were calculated. These correlations tended to be very similar, therefore only the Pearson correlations are reported in the main text. Spearman correlations are reported in the supplementary materials. All correlations carried out for this study, and their exact p-values, are reported in Supplementary Table S8 (Pearson correlations) and Supplementary Table S9 (Spearman rank-order correlations).

### Correlations between cognitive test scores and demographic and health variables

The Pearson correlations between each of the UK Biobank tests and age is shown in Table 2. Note that, UKB RT, UKB TMT part A and UKB TMT part B are measuring response times. On these tests, higher scores indicate that the participants took longer to complete the tests and therefore higher scores reflect poorer performance. The score for UKB Pairs Matching is the number of errors made matching all of the cards and, therefore, a higher score on this test also reflects poorer performance. For all other UK Biobank tests, higher scores indicate better performance. All UK Biobank tests correlated significantly with age. In all but one, older individuals performed more poorly on these tests (absolute *r* = 0.16 to 0.60, *p* ≤ .040). The exception was UKB Picture Vocabulary where older participants performed better than younger participants on this test (*r* = 0.18, *p* = .022). The strongest age associations were seen for tests measuring processing speed. Older adults tended to have lower scores on UKB Symbol Digit (*r* = - 0.60, *p* < .001), and were slower on UKB TMT part A (*r* = 0.58, *p* < .001), and UKB TMT part B (*r* = 0.57, *p* < .001). UKB Pairs Matching, UKB Tower test, and UKB Matrices also had absolute correlations of > 0.3 (*p* < .001) with age.

**Table 2.**
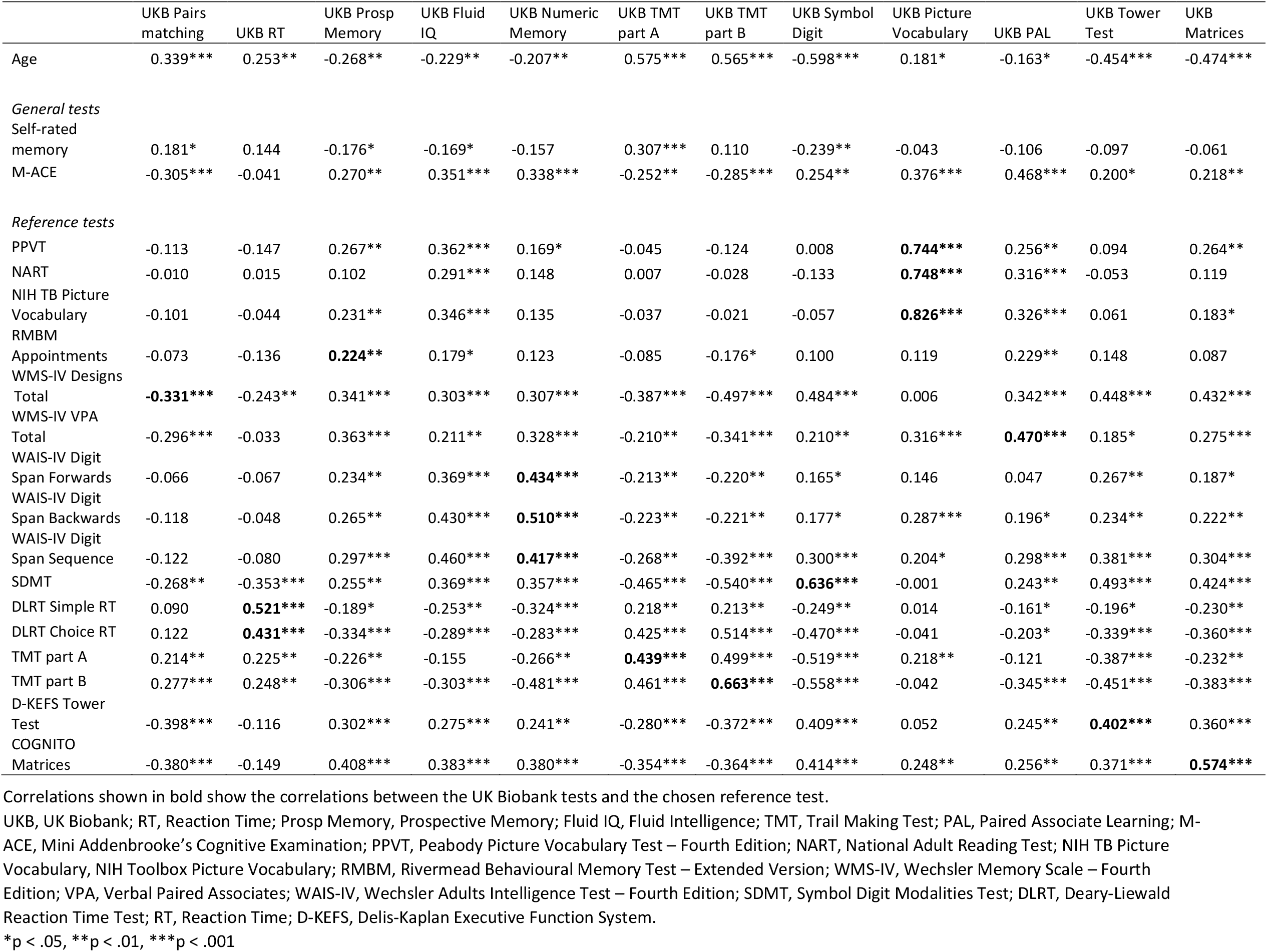
Pearson correlations between UK Biobank tests with age, general tests, and reference tests (n = 154-160)

The Pearson and Spearman rank-order correlations between all cognitive tests and age, sex, years of education, and general health are shown in Supplementary Tables S10 and S11. Male participants had lower scores than female participants on UKB PAL (*r* = −0.28, *p* < .001), but higher scores on the UKB Tower Test (*r* = 0.19, *p* = .018) and UKB Matrices (*r* = 0.17, *p* = .034). Individuals with more years of education were quicker on UKB TMT parts A (*r* = −0.18, *p* = .024) and B (*r* = −0.21, *p* = .007), and scored higher on UKB Picture Vocabulary (*r* = 0.29, *p* < .001) and UKB Matrices (*r* = 0.32, *p* < .001). None of the UK Biobank tests were associated with general health.

#### Associations with general tests

The Pearson correlations between the UK Biobank tests and the general tests are reported in Table 2. The Spearman rank-order correlations are reported in Supplementary Table S12. Reporting poorer self-rated memory was associated with more errors on UKB Pairs Matching (*r* = 0.18, *p* = .022), being less likely to correctly touch the orange circle in UKB Prospective Memory (*r* = −0.18, *p* = .026), having lower scores on UKB Fluid IQ (*r* = −0.17, *p* = .033) and UKB Symbol Digit (*r* = −0.24, *p* = .002), and being slower on UKB TMT part A (*r* = 0.31, *p* < .001).

Except for UKB RT, higher scores on the M-ACE were associated with better performance on all UK Biobank cognitive tests, with absolute effect sizes of 0.2 or higher. The M-ACE was most strongly correlated with performance on UKB PAL (*r* = 0.47, *p* < .001). UK Picture Vocabulary (*r* = 0.38, *p* < .001) also correlated moderately with the M-ACE.

#### Associations with reference tests

The Pearson correlations between the UK Biobank cognitive tests and the reference tests are reported in Table 2. The Spearman rank-order correlations are reported in Supplementary Table S12. The correlations highlighted in bold in Table 2 (and Supplementary Table S12) reflect the correlations between the UK Biobank test and the chosen reference test which was judged to be assessing the same cognitive capability or domain. In going through each UK Biobank cognitive test’s results below we first describe the correlation with the respective reference test(s), and then we highlight some correlations with ‘non-reference’ tests that have absolute effect sizes greater than about 0.3.

### UKB Pairs Matching

More errors on UKB Pairs Matching was associated with higher scores on WMS-IV Designs Total—the reference test for UKB Pairs Matching (*r* = −0.33, *p* < .001). UKB Pairs Matching test was also moderately associated with D-KEFS Tower Test (*r* = −0.40, *p* < .001) and COGNITO Matrices (*r* = −0.38, *p* < .001). D-KEFS Tower Test and COGNITO Matrices are both visuospatial reasoning tests. The correlations between UKB Pairs Matching with D-KEFS Tower Test and COGNITO Matrices suggests this test may be measuring reasoning and executive function, as well as visual memory.

### UKB RT

The DLRT Simple and Choice RT were chosen as reference tests for UKB RT. Slower response on UKB RT was associated with slower DLRT Simple RT (*r* = 0.52, *p* < .001) and DLRT Choice RT (*r* = 0.43, *p* < .001). The SDMT, another measure of processing speed, also correlated moderately with the UKB RT such that individuals who had quicker responses on UKB RT scored higher, and were therefore quicker, on the SDMT (*r* = −0.35, *p* < .001).

### UKB Prospective Memory

There was a small, positive correlation between UKB Prospective Memory and the chosen reference test, the RMBM Appointments (*r* = 0.22, *p* = .005). All other reference tests, except NART and DLRT Simple RT, had stronger correlations with UKB Prospective Memory than that reported between UKB Prospective Memory and RMBM Appointments. Correctly answering the UKB Prospective Memory test on the first attempt was most strongly associated with higher scores on COGNTIO Matrices (*r* = 0.41, *p* < .001) and WMS-IV VPA Total (*r* = 0.36, *p* < .001).

### UKB Fluid IQ

A number of the general and reference tests correlated positively at *r* > 0.30 with UKB Fluid IQ, including M-ACE, PPVT, NIH Toolbox Picture Vocabulary, WMS-IV Designs Total, WAIS-IV Digit Span Forwards, Backwards, and Sequence, SDMT, TMT part B, and COGNITO Matrices. The strongest correlations were for WAIS-IV Digit Span Backwards (*r* = 0.43, *p* < .001), WAIS-IV Digit Span Sequence (*r* = 0.46, *p* < .001), and COGNITO Matrices (*r* = 0.38, *p* < .001), which are tests designed to assess working memory and reasoning, i.e. cognitive domains which are thought to be fluid abilities.

### UKB Numeric Memory

This test correlated positively with the chosen reference test—WAIS-IV Digit Span. The correlations for WAIS-IV Digit Span Forwards, Backwards and Sequence were 0.43, 0.51, and 0.42, respectively (for all, *p* < .001). UKB Numeric Memory also correlated moderately with TMT part B, such that individuals who scored higher on UKB Numeric Memory were quicker on TMT part B (*r* = - 0.48, *p* < .001). UKB Numeric Memory also had correlations > 0.3 with M-ACE, WMS-IV Designs Total, WMS-IV VPA Total, SDMT, DLRT Simple RT, and COGNITO Matrices.

### UKB TMT part A

UKB TMT part A correlated positively with TMT part A (*r* = 0.44, *p* < .001)—the chosen reference test—and TMT part B (*r* = 0.46, *p* < .001). UKB TMT A also correlated highly with the SDMT, another measure of processing speed, such that individuals who scored higher on the SDMT were quicker on UKB TMT A (*r* = −0.47, *p* < .001).

### UKB TMT part B

UKB TMT part B was strongly and positively correlated with the paper-and-pencil version of the TMT part B (the reference test; *r* = 0.66, *p* < .001), and also with TMT part A (*r* = 0.50, *p* < .001). Being quicker on UKB TMT part B was also highly correlated with being quicker on DLRT Choice RT (*r* = 0.54, *p* < .001) and having higher scores on SDMT (*r* = −0.54, *p* < .001); both are tests of processing speed. Both TMT parts A and B had several other moderately strong correlations with other tests.

### UKB Symbol Digit

Higher scores on UKB Symbol Digit were correlated at *r* = 0.64 (*p* < .001) with higher scores on the SDMT, the reference test for UKB Symbol Digit. Better performance on this test also correlated at > 0.5 with being quicker at TMT part A (*r* = −0.52, *p* < .001) and part B (*r* = −0.56, *p* < .001), both of which have a speed component. There were also correlations > 0.4 with WMS-IV Designs Total, DLRT Choice RT, D-KEFS Tower Test, and COGNITO Matrices.

### UKB Picture Vocabulary

UKB Picture Vocabulary correlated highly with its three reference tests: 0.83 with the NIH Toolbox Picture Vocabulary, 0.75 with the NART, and 0.74 with PPVT (for all, *p* < .001). Higher scores on UKB Picture Vocabulary were associated with higher scores on the M-ACE (*r* = 0.38, *p* < .001) and WMS-IV VPA Total score (*r* = 0.32, *p* < .001). These tests have verbal contents.

### UKB PAL

UKB PAL was positively correlated with the WMS-IV VPA Total, the reference test for UKB PAL (*r* = 0.47, *p* < .001). There was also a moderate positive correlation between the UKB PAL and M-ACE (*r* = 0.47, *p* < .001), which includes a verbal memory component (e.g., remembering a name and address). UKB PAL had correlations > 0.3 with NART, NIH Picture Vocabulary, WMS-IV Designs Total, and TMT part B.

### UKB Tower Test

UKB Tower Test correlated positively with its reference test, the D-KEFS Tower Test (*r* = 0.40, *p* < .001). Better performance on the UKB Tower Test was associated with better performance on SDMT (*r* = 0.49, *p* < .001), TMT part B (*r* = −0.45, *p* < .001), and WMS-IV Designs Total (*r* = 0.45, *p* < .001). The correlation with SDMT may reflect the fact that the UKB Tower Test was a timed test— participants were tasked with completing as many Tower trials as possible in 3 minutes—and may therefore be measuring processing speed as well as executive function. The WMS-IV Designs is a measure of visuospatial memory. The UKB Tower Test requires participants to mentally move the hoops on the pegs in their mind, therefore it is likely to also be measuring visuospatial abilities. UKB Tower Test also correlated at > 0.3 with WAIS-IV Digit Span Sequence, DLRT Choice RT, TMT part A, and COGNITO Matrices.

### UKB Matrices

This test correlated positively at *r* = 0.57 (*p* < .001) with the original version of this test; the COGNITO Matrices. Performance on this test also correlated moderately and positively with SDMT (*r* = 0.42, *p* < .001), WMS-IV Designs Total (*r* = 0.43, *p* < .001), WAIS-IV Digit Span Sequence (*r* = 0.03, *p* < .001), DLRT Choice RT (*r* = −0.36, *p* < .001), TMT part B (*r* = −0.38, *p* < .001), and D-KEFS Tower Test (*r* = 0.36, *p* < .001); these are tests that were designed to assess cognitive domains thought to be fluid.

#### Partial correlations adjusting for age

The age-adjusted correlations between the UK Biobank tests and the general and reference tests are reported in Supplementary Table S13 (Pearson correlations) and Supplementary Table S14 (Spearman correlations). After controlling for age, none of the correlations between self-rated memory and the UK Biobank tests were significant. The exception was the correlation between self-rated memory and UKB TMT part A, which remained significant, though reduced in size (*r* = 0.31; age-adjusted *r* = 0.17). All correlations between M-ACE and the UK Biobank tests were smaller—though most remained significant—when adjusting for age, except for the correlation between M-ACE and UKB Picture Vocabulary, which became stronger (*r* = 0.38; age-adjusted *r* = 0.43). The correlations between UKB Tower Test (*r* = 0.20; age-adjusted *r* = 0.12) and UKB Matrices (*r* = 0.22; age-adjusted *r* = 0.14) with the M-ACE were no longer significant when adjusting for age.

Generally, the age-adjusted correlations between the UK Biobank tests and the reference tests tended to be smaller than the raw correlations, though the difference between the raw correlations and the age-adjusted correlations was small. The largest differences were seen for the correlations between the following UK Biobank tests with their respective reference tests: UKB TMT part B (*r* = 0.66; age-adjusted *r* = 0.55), UKB Symbol Digit (*r* = 0.64; age-adjusted *r* = 0.45), UKB Pairs Matching (*r* = −0.33; age-adjusted *r* = −0.19), and UKB TMT part A (*r* = 0.44; age-adjusted *r* = 0.24). For all other correlations between UK Biobank cognitive tests and the reference tests, the change in the strength of the correlation between the raw correlations and the age-adjusted correlations was ≤ 0.07.

#### Measures of general cognitive ability

The correlation between a measure of general cognitive ability created using 11 well-validated reference tests (*g:reference-11*) and 11 UK Biobank tests (*g:UKB-11*) was *r* = 0.83 (*p* < .001; age-adjusted *r* = 0.79, *p* < .001). The correlation between *g:reference-11* and a measure of general ability created using the five UK Biobank baseline tests (*g:UKB-5*) was *r* = 0.74 (*p* < .001; age-adjusted *r* = 0.69, *p* < .001).

### Sensitivity analysis

The UKB Picture Vocabulary and UKB Matrices tests shared items with the NIH Toolbox Picture Vocabulary and the COGNITO Matrices tests. To check whether the high correlation found between *g:reference-11* and *g:UKB-11* was because of this overlap in items, another measure of general cognitive ability was created using the same tests as in *g:reference-11*, but excluding scores on the COGNITO Matrices and NIH Toolbox Picture Vocabulary (*g:reference-9*). The results of a PCA using 9 reference tests are reported in the Supplementary materials (scree plot of the eigenvalues is shown in Supplementary Fig. S5; the results of the PCA are reported in Supplementary Table S15). The correlations between *g:reference-9* and *g:UKB-11* (*r* = 0.85, *p* < .001; age-adjusted *r* = 0.78, *p* < .001) and between *g:reference-9* and *g:UKB-5* (*r* = 0.73, *p* < .001; age-adjusted *r* = 0.67, *p* < .001) were similar in size to that found when using *g:reference-11.*

### Correlations between g:reference-11 and UK Biobank tests

Correlations and age-adjusted correlations between *g:reference-11* and each of the UK Biobank tests are reported in Supplementary Table S16 (Pearson correlations) and Supplementary Table S17 (Spearman rank-order correlations). All UK Biobank cognitive tests correlated with this measure of general cognitive ability, such that higher scores on *g:reference-11* were associated with better performance on the UK Biobank cognitive tests (*p* < .001). UKB RT had the lowest correlation with *g:reference-11* (*r* = −0.29, *p* <.001), whereas UKB TMT part B had the strongest correlation (*r* = −0.62, *p* < .001). Other UK Biobank tests which correlated positively with general cognitive ability at > 0.5 were UKB Fluid IQ (*r* = 0.55, *p* < .001), UKB Numeric memory (*r* = 0.55, *p* < .001), UKB Symbol Digit (*r* = 0.54, *p* < .001), UKB Tower Test (*r* = 0.52, *p* < .001) and UKB Matrices (*r* = 0.58, *p* < .001). This makes the important point that, although each individual test is often associated with a specific cognitive domain, all of the tests are loaded, to different extents, on general cognitive ability.

The age-adjusted Pearson correlations between *g:reference-11* and UK Biobank tests (Supplementary Table S16) tended to be weaker than the raw correlations, except for the correlation between *g:reference-11* and UKB Picture Vocabulary, which became stronger (raw *r* = 0.43, *p* < .001; age-adjusted *r* = 0.58, *p* <.001).

### Correlations between g:UKB-11 and g:UKB-5 with the general and reference tests

The correlations and age-adjusted correlations between general cognitive ability created using the UK Biobank tests (*g:UKB-11* and *g:UKB-5*) and the general tests and reference tests are shown in Supplementary Tables S18 (Pearson correlations) and S19 (Spearman rank-order correlations). Higher scores on *g:UKB-11* were associated with better performance on all the general and reference tests, except for self-rated memory and the NART which were not significantly associated with *g:UKB-11*. Higher *g:UKB-11* score was most strongly related to better performance on SDMT (*r* = 0.68, *p* < .001), TMT part B (*r* = −0.64, *p* < .001), and WMS-IV Designs Total (*r* = 0.63, *p* < .001), suggesting that the general cognitive ability measure created using all UK Biobank tests is influenced by speed of processing.

When adjusting for age, some of the associations between *g:UKB-11* and the reference tests reduced in strength (e.g., the correlation with tests of speed, executive function, and reasoning), whereas others became stronger (e.g., correlations with vocabulary tests, RMBM Appointments, and WAIS-IV Digit Span). However, when adjusting for age, all test except self-rated memory were associated with *g:UKB-11* such that a better *g:UKB-11* score was associated with better test scores on the general and reference tests. Whereas there was no association between *g:UKB-11* and the NART when calculating raw correlations (*r* = 0.10, *p* > .05), there was a moderate and positive association between *g:UKB-11* and the NART when adjusting for age (age-adjusted *r* = 0.35, *p* < .001).

Higher scores on *g:UKB-5* were also associated with better performance on the general and reference tests, except the NART, which was not associated with *g:UKB-5*. Again, the association between *g:UKB-5* and the NART became significant when adjusting for age (*r* = 0.31, *p* < .001). Generally, the correlations seen between *g:UKB-5* and the reference tests were lower than those seen between *g:UKB-11* and the reference tests.

#### UK Biobank questionnaire

### Clear test instructions

The number and percentage of participants who thought the UK Biobank tests were unclear is reported in Supplementary Table S20. A total of 8 (5.5%) participants reported that they thought the UK Biobank test instructions in general were unclear. Nearly one quarter of participants (n = 35, 24.1%) reported that they thought the instructions for the UKB Tower Test were not clear. Participants generally thought the instructions for UKB RT, UKB Picture Vocabulary and UKB Matrices were clear. Only 3 (2.1%), 2 (1.4%), and 1 (0.7%) participants, respectively, reported that the instructions for these tests were not clear.

### UKB Numeric Memory technique

Of the 141 individuals who were asked about the technique used to complete the Numeric memory test, only 20 (14.2%) participants reported that they performed the UKB Numeric Memory test as a backward digit span (e.g., read from left-to-right and reversed the digits in their mind). Most (n = 102; 72.3%) performed a forward digit span (e.g., read from right-to-left and did not reverse the digits in their mind). The remaining participants (n = 19; 13.5%) reported using a mixture of both techniques. Participants who did a backward digit span (mean = 6.70, SD = 1.08) had a slightly lower mean score on UKB Numeric Memory than those who did a forward digit span (mean = 6.92, SD = 1.24) or those who did a mixture of both (mean = 7.11, SD = 1.25). However, a between-group ANOVA revealed that UKB Numeric Memory scores did not differ by technique used (F (2, 136) = 0.518, *p* = .602).

#### Test-retest reliability

Table 3 reports the means and SDs for all of the UK Biobank tests at Time 1 and Time 2 in a subsample of participants (n = 52) who completed the UK Biobank tests for a second time after a mean interval of 28.88 days (SD = 2.02). Generally, mean scores were higher at Time 2 than at Time 1; however, mean performance was significantly better at Time 2 compared to Time 1 only for UKB RT (*t*(51) = 3.22, *p* = .002), UKB Fluid IQ (*t*(50) = 2.26, *p* = 0.03), and UKB Symbol Digit (*t*(51) = 3.37, *p* = .001). The effect size for the difference between mean score at Time 1 and Time 2 ranged from negligible (Cohen’s *d* = 0.00) to moderate (Cohen’s *d* = 0.47). UKB Fluid IQ (Cohen’s *d* = 0.32), UKB RT (Cohen’s *d* = 0.45) and UKB Symbol Digit (Cohen’s *d* = 0.47) may be the most prone to repeat testing effects.

**Table 3.** Test-retest^a^ reliability for the UK Biobank cognitive tests (n = 52) and comparable reference tests

| UK Biobank tests | Time 1 |  | Time 2 |  | *p* | Cohen's *d* | Pearson *r*<sub>12</sub> | Spearman *rho*<sub>12</sub> | Reference tests |  |
| --- | --- | --- | --- | --- | --- | --- | --- | --- | --- | --- |
|  | Mean | SD | Mean | SD |  |  |  |  | Reference test | Reference test *r*<sub>12</sub> |
| UKB Pairs Matching | 3.08 | 2.63 | 2.79 | 2.07 | 0.427 | 0.11 | 0.409** | 0.348* | WMS-IV Designs I<sup>d</sup> | 0.73 |
|  |  |  |  |  |  |  |  |  | WMS-IV Designs II<sup>d</sup> | 0.72 |
| UKB Reaction time | 627.04 | 98.37 | 588.87 | 77.50 | 0.002 | 0.45 | 0.550*** | 0.585*** | DLRT Simple RT<sup>e</sup> | 0.64 |
|  |  |  |  |  |  |  |  |  | DLRT Choice RT<sup>e</sup> | 0.83 |
| UKB Prospective Memory | 37 | 71.2% | 48 | 92.3% | - | 41 (78.8%)<sup>c</sup> | 0.453** | 0.453** | - | - |
| UKB Fluid IQ<sup>b</sup> | 6.80 | 2.05 | 7.39 | 2.14 | 0.028 | 0.32 | 0.607*** | 0.560*** | - | - |
| UKB Numeric Memory | 7.00 | 1.24 | 7.00 | 1.17 | 1.000 | 0.00 | 0.501*** | 0.517*** | WAIS-IV Digit Span<sup>f</sup> | 0.82 |
| UKB TMT part A | 246.85 | 90.59 | 231.98 | 72.22 | 0.211 | 0.18 | 0.478*** | 0.502*** | TMT part A<sup>g</sup> | 0.79 |
| UKB TMT part B | 475.52 | 137.51 | 466.42 | 151.98 | 0.506 | 0.09 | 0.775*** | 0.861*** | TMT part B<sup>g</sup> | 0.89 |
| UKB Symbol Digit | 18.85 | 5.63 | 21.12 | 4.91 | 0.001 | 0.47 | 0.583*** | 0.603*** | SDMT<sup>h</sup> | 0.80 |
| UKB Picture Vocabulary | 8.94 | 1.59 | 9.08 | 1.55 | 0.166 | 0.19 | 0.887*** | 0.828*** | NIH TB Picture Vocabulary<sup>i</sup> | ICC = 0.81 (95% CI 0.73 to 0.87) |
|  |  |  |  |  |  |  |  |  | PPVT<sup>j</sup> | 0.94 |
| UKB PAL | 7.87 | 2.22 | 8.33 | 2.20 | 0.158 | 0.20 | 0.450** | 0.542*** | WMS-IV VPA I<sup>d</sup> | 0.79 |
|  |  |  |  |  |  |  |  |  | WMS-IV VPA II<sup>d</sup> | 0.81 |
| UKB Tower Test | 10.48 | 3.36 | 11.10 | 3.07 | 0.202 | 0.18 | 0.434** | 0.410** | D-KEFS Tower test<sup>k</sup> | 0.44 |
| UKB Matrices | 9.02 | 1.79 | 9.40 | 1.84 | 0.153 | 0.20 | 0.445** | 0.442** | COGNITO Matrices<sup>l</sup> | 0.70 |

<sup>a</sup>Test-retest interval = 28.88 (SD = 2.02) days.<sup>b</sup>n = 51<sup>c</sup>For UKB Prospective memory, the value is n (percentage) agreement for whether participants gave the same response (i.e., correct or incorrect on first attempt) at Time 1 and Time 2.<sup>d</sup>Test-retest interval mean = 23 days (range 14 to 84 days), n = 244<sup>11</sup>.<sup>e</sup>Period-free reliability. Participants completed the DLRT Simple RT and DLRT Choice RT twice immediately one after the other, n = 20<sup>19</sup>.<sup>f</sup>Test-retest interval mean = 22 days (range 8 to 82 days), n = 298<sup>33</sup>.<sup>g</sup>Test-retest interval mean = 11 months, n = 384<sup>23</sup>.<sup>h</sup>Test-retest interval mean = 29.40 days, n = 80<sup>12</sup>.<sup>i</sup>Test-retest interval range 7 to 21 days, n = 89<sup>15</sup>.
<sup>j</sup>Test-retest interval mean = 30.1 days (range 14 to 48 days), n = 67<sup>22</sup>.
<sup>k</sup>Test-retest interval mean = 25 days (range 9 to 74 days), n = 101<sup>13</sup>.
<sup>l</sup>Test-retest interval range 2 to 3 weeks, n = 78<sup>24</sup>.
*p*, *p*-value from a paired-samples t-test comparing scores at Time 1 and Time 2; *r*<sub>12</sub>, Pearson correlation between test scores at Time 1 and Time 2; *rho*<sub>12</sub>, Spearman rank-order correlation between test scores at Time 1 and Time 2; UKB, UK Biobank; Fluid IQ, Fluid Intelligence; TMT, Trail Making Test; PAL, Paired Associate Learning; Matrices, Matrix Pattern Test; WMS-IV, Wechsler Memory Scale – Fourth Edition; DLRT, Deary-Liewald Reaction Time Test; RT, Reaction Time; WAIS-IV, Wechsler Adult Intelligence Scale – Fourth Edition; SDMT, Symbol Digit Modalities Test; NIH TB Picture Vocabulary, NIH Toolbox Picture Vocabulary Test; PPVT, Peabody Picture Vocabulary Test – Fourth Edition; VPA, Verbal Paired Associates; D-KEFS, Delis-Kaplan Executive Function System; ICC, Intraclass correlation.
\**p* < .05, \*\**p* < .01, \*\*\**p* < .001

The test-retest reliabilities (Pearson and Spearman rank-order correlations) for each UK Biobank test are reported in Table 3. This table also contains some of the test-retest correlations reported elsewhere for some of the reference tests. UKB Pairs Matching had the lowest Pearson test-retest correlation (*r*_12_ = 0.41, *p* = .003). Test-retest reliability was high for UKB Picture Vocabulary (*r*_12_ = 0.89, *p* < .001) and UKB TMT B (*r*_12_ = 0.78, *p* < .001). Test-retest correlations for all other UK Biobank tests were moderate-to-high (*r*_12_ = 0.43 to 0.61). The test-retest reliability found here for the UK Biobank tests tended to be lower than those reported elsewhere for the reference tests. For example, the test-retest reliability for WMS-IV VPA I and VPA II was *r*_12_ = 0.79 and *r*_12_ = 0.81, respectively^11^, whereas the test-retest correlation for UKB PAL was *r*_12_ = 0.45. The SDMT has been reported to have a test-retest reliability of 0.80 over a 30 day test-retest interval^12^, whereas the test-retest reliability here for UKB Symbol Digit was found to be *r*_12_ = 0.58. Whereas the test-retest reliability reported here for the UKB Tower Test was relatively low (*r*_12_ = 0.43), this test-retest correlation was in line with that previously reported for the D-KEFS Tower Test (*r*_12_ = 0.44^13^).

## Discussion

Using a sample of 160 middle-aged and older adults this study investigated the concurrent validity and test-retest reliability of the UK Biobank cognitive tests. This study had three main findings: 1) generally, the UK Biobank tests correlated moderately-to-strongly with well-validated, standard tests designed to assess the same cognitive domain; 2) a measure of general cognitive ability can be created using all of the UK Biobank tests, as well as using only the five UK Biobank baseline tests, and these measures of general cognitive ability are highly correlated with a measure of general cognitive ability created using a battery of standard cognitive measures; 3) most of the UK Biobank tests showed moderate-to-high test-retest reliability, but these tended to be lower than those reported elsewhere for the reference tests.

### Concurrent validity

Prior to discussing this part of the study’s results, we acknowledge that most tests load strongly on the general cognitive component. Therefore, we are aware that, when we write about tests correlating because they both assess the same ‘cognitive domain’ or ‘underlying cognitive ability’—especially with respect to UK Biobank tests versus their reference tests—it might also be in part or in whole because they both assess general cognitive ability (*g*). It is an error not to acknowledge this, as Schmidt^14^ discusses in detail. However, mindful of the fact that there is variance beyond *g* accounted for at the level of cognitive domains and specific abilities^9^, and the fact that readers will wish to know how the largely-undocumented UK Biobank tests relate to better-validated tests, we think the references we make to domains and specific abilities are appropriate.

Despite the brief, and non-standard nature of the UK Biobank cognitive assessment, the UK Biobank tests tended to correlate moderately-to-strongly with well-validated cognitive tests that were designed to assess the same cognitive domain or specific ability. Thus, the UK Biobank cognitive tests mostly showed modest to good concurrent validity. The UK Biobank Picture Vocabulary test showed especially good concurrent validity. This test correlated very highly (*r* = 0.83) with the original version of this test—the NIH Toolbox Picture Vocabulary test—and also with another picture vocabulary test, the PPVT (*r* = 0.74). In a validation study of the NIH Toolbox^15^, the correlation between the NIH Toolbox Picture Vocabulary test and the PPVT was *r* = 0.78, which is very similar to the correlation found here between the UK Biobank version of this test and the PPVT (*r* = 0.74). In addition, the UKB Picture Vocabulary test was also found to correlate highly (*r* = 0.75) with the NART, which is often used as an estimate of crystallised cognitive ability^16,17^. The results from this study suggest that the UK Biobank Picture Vocabulary is a valid measure of crystallised ability and may be used as an estimate of premorbid cognitive functioning.

Other tests with good concurrent validity are the UKB TMT part B and UKB Symbol Digit tests, which both correlated at greater than 0.6 with the original, paper-and-pencil versions of these tests^12,18^. The UKB RT score, which is created from a mean of only 4 trials, correlated at 0.52 with DLRT Simple RT and at 0.43 with DLRT Choice RT, which are more detailed tests of reaction time created from a mean of 20 trials and 40 trials, respectively^19^.

A previous study using UK Biobank baseline data^20^ found that scores on the UKB Fluid IQ test showed mean values that remained relatively stable between the ages of 40 and 60 years and therefore did not show the age-related decline across the adult lifespan that is the hallmark of fluid ability^2^. Hagenaars et al.^20^ suggested that, because of the relative stability in middle-age, UKB Fluid IQ may in fact be measuring a more crystallised ability. In the present study, we did not include a reference test for UKB Fluid IQ because we were unable to identify an appropriately similar test. However, we found that UKB Fluid IQ was negatively correlated with age and that it correlated most strongly (*r* ≥ 0.38) with tests of working memory (WAIS-IV Digit Span), and non-verbal reasoning (COGNTO Matrices)— tests thought to assess more fluid abilities^2^. Therefore, this test may be more fluid than was suggested by Hagenaars et al.^20^, although it did also have moderate correlations with the three standard vocabulary tests.

The UKB PAL test exhibited negative skew (Supplementary Fig. S1) suggesting that most participants find this test quite easy. Despite the negative skew, scores on the UKB PAL test were found to correlate moderately with the M-ACE (*r* = 0.47), a brief assessment of global cognitive functioning that is designed to identify individuals who may have possible cognitive impairment^21^. The UKB PAL test may be a useful test to identify individuals in UK Biobank who may have a possible cognitive impairment.

### General cognitive ability

Previous research has found that measures of general cognitive ability (*g*) created using entirely different sets of cognitive tests correlate very strongly^5,6^. In the present study, we compared whether measures of general cognitive ability created using the brief, bespoke UK Biobank tests that were administered unsupervised correlated strongly with a measure of general cognitive ability created using well-validated tests administered under standardised conditions. The correlations between general cognitive ability created using well-validated tests and general cognitive ability created using the UK Biobank tests were high (*r* = 0.83 for a measure created using all 11 UK Biobank tests; *r* = 0.74 for the 5 baseline UK Biobank tests). These correlations were lower than those reported in one study^5^ that found that three measures of general cognitive ability created using three entirely different cognitive test batteries correlated nearly perfectly (*r* ≥ 0.99). However, the correlations found here are in line with another study^6^ that compared five different measures of general cognitive ability and found that they correlated at *r* ≥ 0.77. This suggests that, despite the brief and non-standard nature of the UK Biobank cognitive assessment, a measure of general cognitive ability can be created using these tests. UKB TMT part B, UKB Matrices, UKB Numeric Memory, and UKB Fluid IQ all correlated at ≥ 0.55 with the general measure of cognitive ability created using the standardised tests, suggesting these UK Biobank tests load strongly on general cognitive ability.

### Reliability

Test-retest correlations for UKB Picture Vocabulary (*r*_12_ = 0.89) and UKB Trail Making Test part B (*r*_12_ = 0.78) were high, and comparable to those reported for other, well-validated, measures of picture vocabulary (NIH Toolbox Picture Vocabulary intraclass correlation = 0.81; 95% CI 0.73 to 0.87^15^; and PPVT *r*_12_ = 0.94^22^) and for the original paper-and-pencil version of the TMT part B (*r*_12_ = 0.89^23^). Therefore, the UK Biobank Picture Vocabulary test and TMT part B show good stability. Good short-term stability of cognitive tests is especially important when examining longitudinal change. Low stability means that any differences in scores over time may not be due to real change in test performance, but due to error of measurement. Generally, the test-retest reliability for the UK Biobank tests was substantial; however, they tended to be lower than those reported for the reference tests, suggesting that the UK Biobank tests are less stable across time than well-validated tests administered under standardised conditions. The relative brevity of some of the UK Biobank tests might contribute to the lower reliability. However, UKB Matrices uses the same 15 items as COGNITO Matrices, and both are administered via a computer and yet the test-retest correlation for UKB Matrices was *r*_12_ = 0.44, whereas the test-retest correlation reported for the COGNTIO Matrices test was higher, at *r*_12_ = 0.70^24^. It is not clear why the UK Biobank tests have lower test-retest reliability than other measures of cognitive function.

UKB Pairs Matching had the lowest test-retest reliability (*r*_12_ = 0.41). However, the test-retest reliability reported in this study for UKB Pairs Matching was substantially larger than has previously been reported for this test using UK Biobank data^3,20^. Lyall et al.^3^ calculated the correlation between performance on the UKB Pairs Matching test at baseline and at the repeat study approximately four years later and reported a test-retest reliability of *r*_12_ = 0.19^3^. The lower test-retest reliability reported in Lyall et al.^3^ might be because they used a test-retest interval of over 4 years, which is much longer than the conventional 2-4 week test-retest interval. Lyall et al.^3^ reported relatively similar test-retest correlations for UKB RT (Lyall et al. *r*_12_ = 0.54^3^; present study *r*_12_ = 0. 55) and UKB Fluid IQ (Lyall et al. *r*_12_ = 0.65^3^; present study *r*_12_ = 0.61) to that reported this study, despite the different test-retest intervals.

### Other psychometric considerations in some UK Biobank cognitive tests

The UKB Numeric Memory test was designed as a backwards digit span task to assess working memory—the ability to temporarily store information in short-term memory long enough to manipulate it^17^. Backward digit span tasks require individuals to both remember a sequence of numbers and mentally reverse these numbers in their mind, and this differs from a forward digit span task where participants are only required to remember a sequence of digits^17^. Despite the fact that the UKB Numeric Memory test was designed to assess backward digit span, we found that only 14.2% of the sample tested in the current study reported performing a backward digit span. All other participants reported that they either carried out a forward digit span (72.3%), or they used a mixture of both techniques (13.5%). This means that, for the majority of participants, this test is not assessing the type of mental performance that it was intended to assess.

This study also found that nearly one-quarter of participants reported that they thought the test instructions for the UKB Tower Test were unclear. Given that the UK Biobank tests are administered unsupervised, and participants are expected to sit at a computer in a UK Biobank clinic and work through these tests independently, it is important that the test instructions are clear and the participant knows exactly what to do before starting the test proper. The UKB Tower Test had several pages of instructions. The length of the test instructions might be an important contributor to why participants reported that the test instructions for UKB Tower Test were unclear. Other UK Biobank tests with lengthy instructions, including UKB Symbol Digit (9.7%) and UKB TMT (8.3%) also had higher percentages of participants reporting that the test instructions for these tests were not clear, whereas test with relatively short instructions, such as UKB Matrices (0.7%) and UKB Picture Vocabulary (1.4%) tended to have very few participants reporting that they thought the instructions were not clear. All of these tests, however, had practice examples which will have allowed participants to see what is involved before starting the task proper, even if they did not fully understand the test instructions before starting the practice trials.

### Advantages and limitations

The main advantage of this study is that the fully-automated UK Biobank cognitive assessment was compared to a large number of well-validated, standard cognitive tests that were administered under standardised conditions. This meant that the brief and non-standard UK Biobank cognitive tests were compared to what many would consider to be the ‘gold standard’ measures of cognitive ability. For the current study, UK Biobank programmed a stand-alone version of the UK Biobank cognitive assessment that is currently being administered at the UK Biobank imaging study. UK Biobank also provided us with a UK Biobank button-box to be used for the UKB RT test and with details about the computing equipment used at the UK Biobank clinic assessments which enabled us to very closely mimic the UK Biobank clinic cognitive assessment.

However, there are some limitations to the current study. The sample size is relatively small, especially for the test-retest sample. The testing conditions in the current study were not identical to the testing conditions used during the UK Biobank clinic assessments. In the current study, participants were assessed individually in a quiet room, free of distraction. The UK Biobank assessment centre could be busy and sometimes noisy (CF-R and IJD both spent a day at one UK Biobank testing centre during people’s imaging visits). Therefore, in the current study, the UK Biobank tests were administered in a more usual and standardised psychological testing environment. It is not clear whether the reliability and validity reported in the current study would differ if the UK Biobank tests had been administered in a busy and sometimes noisy environment that was seen when the authors visited the UK Biobank testing centre. In addition to the cognitive assessment administered at the UK Biobank assessment centre, UK Biobank have also collected cognitive data using a web-based assessment. For the web-based assessment, participants are sent a link, via email, and were to complete the cognitive tests at home. The testing conditions of the web-based assessment, therefore, were even less controlled than at the UK Biobank assessment centre. We do not know whether the results of the current study would generalise to the UK Biobank web-based assessment.

## Conclusions

This study examined the concurrent validity and test-retest reliability of the enhanced UK Biobank cognitive assessment that is currently being administered to UK Biobank participants attending the UK Biobank imaging study. The UK Biobank cognitive tests are administered using a fully-automated touch-screen assessment, and participants complete these tests unsupervised. The tests in UK Biobank tend to be short. They were created specifically for UK Biobank, or were adapted for use in a fully-automated assessment. The present study found that they showed a range of concurrent validity coefficients with well-validated, standard tests of cognitive ability, and they tended to have moderate-to-good test-retest reliability. UK Biobank is one of the largest and most detailed health resources available worldwide. This paper provides currently-lacking information on the psychometric properties of the UK Biobank cognitive tests.

## Data Availability

The dataset generated during the current study are available from the authors on reasonable request.

## Acknowledgements

We thank the participants of the present study. We thank UK Biobank for providing us with a stand-alone version of the UK Biobank cognitive assessment, a UK Biobank button-box, and advice on UK Biobank computer equipment. We especially thank Alan Young, Keith Anderson and other members of the NDPH Core Programming and Testing teams for producing a stand-alone version of the UK Biobank cognitive tests. We thank Michelle Miranthi for her assistance with the data entry. We thank John Gallacher, Marcus Richards, and our late colleague John Starr, who were members of Work Package 10 of Dementias Platform UK (DPUK) alongside IJD and CF-R.

Join Dementia Research was used to identify potential participants for this study. Join Dementia Research is funded by the Department of Health and delivered by the National Institute for Health Research in partnership with Alzheimer Scotland, Alzheimer’s Research UK and Alzheimer’s Society. This work was supported by the University of Edinburgh Centre for Cognitive Ageing and Cognitive Epidemiology, part of the cross council Lifelong Health and Wellbeing Initiative, funded by the Biotechnology and Biological Sciences Research Council (BBSRC), and Medical Research Council (MRC) (grant number MR/K026992/1). CF-R and IJD were supported by DPUK, funded through the MRC (MR/L023784/2).

## Author Contributions

CF-R and IJD contributed to the conception and design of this project. C-FR collected the data. CF-R analysed the data, supervised by IJD. CF-R and IJD wrote and approved the manuscript.

## Additional Information

Competing Interests: IJD is a participant in UK Biobank. IJD declares no other potential conflict of interest. CF-R declares no potential conflict of interest.

## References

1 Sudlow, C. et al. UK biobank: an open access resource for identifying the causes of a wide range of complex diseases of middle and old age. PLoS Medicine 12, e1001779, (2015).

2 Salthouse, T. A. Selective review of cognitive aging. Journal of the International Neuropsychological Society 16, 754–760, (2010).

3 Lyall, D. M. et al. Cognitive Test Scores in UK Biobank: Data Reduction in 480,416 Participants and Longitudinal Stability in 20,346 Participants. PLoS One 11, e0154222, (2016).

4 Spearman, C. “General Intelligence,” objectively determined and measured. Am J Psychol 15, 201–292, (1904).

5 Johnson, W., Bouchard Jr, T. J., Krueger, R. F., McGue, M. & Gottesman, I. I. Just one g: Consistent results from three test batteries. Intelligence 32, 95–107, (2004).

6 Johnson, W., te Nijenhuis, J. & Bouchard Jr, T. J. Still just 1 g: Consistent results from five test batteries. Intelligence 36, 81–95, (2008).

7 Carroll, J. B. Human Cognitive Abilities: A Survey of Factor-Analytic Studies. (Cambridge University Press, 1993).

8 Deary, I. J., Harris, S. E. & Hill, W. D. What genome-wide association studies reveal about the association between intelligence and physical health, illness, and mortality. Curr Opin Psychol 27, 6–12, (2019).

9 Deary, I. J. Intelligence. Current Biology 23, R673–676, (2013).

10 Hsieh, S., Schubert, S., Hoon, C., Mioshi, E. & Hodges, J. R. Validation of the Addenbrooke’s Cognitive Examination III in frontotemporal dementia and Alzheimer’s disease. Dement Geriatr Cogn Disord 36, 242–250, (2013).

11 Wechsler, D. WMS-IV Technical and Interpretive Manual (Pearson, 2009).

12 Smith, A. Symbol Digit Modalities Test: Manual. (Western Psychological Services, 1973).

13 Delis, D. C., Kaplan, E. & Kramer, J. H. D-KEFS Technical Manual. (Pearson, 2001).

14 Schmidt, F. L. Beyond questionable research methods: The role of omitted relevant research in the credibility of research. Arch Sci Psychol 5, 32, (2017).

15 Weintraub, S. et al. Cognition assessment using the NIH Toolbox. Neurology 80, S54–64, (2013).

16 Lara, J. et al. A proposed panel of biomarkers of healthy ageing. BMC Medicine 13, 222, (2015).

17 Mathers, J. C. et al. Guidelines for biomarkers of healthy ageing. (Medical Research Council., 2015).

18 Reitan, R. M. & Wolfson, D. The Halstead-Reitan Neuropsychological Test Battery. (Neuropsychology Press, 1985).

19 Deary, I. J., Liewald, D. & Nissan, J. A free, easy-to-use, computer-based simple and four-choice reaction time programme: the Deary-Liewald reaction time task. Behav Res Methods 43, 258–268, (2011).

20 Hagenaars, S. P. et al. Shared genetic aetiology between cognitive functions and physical and mental health in UK Biobank (N=112 151) and 24 GWAS consortia. Mol Psychiatry 21, 1624–1632, (2016).

21 Hsieh, S. et al. The Mini-Addenbrooke’s Cognitive Examination: a new assessment tool for dementia. Dement Geriatr Cogn Disord 39, 1–11, (2015).

22 Dunn, L. M. & Dunn, D. M. Peabody Picture Vocabulary Test, Fourth Edition: Manual. (Pearson, 2007).

23 Dikmen, S. S., Heaton, R. K., Grant, I. & Temkin, N. R. Test-retest reliability and practice effects of expanded Halstead-Reitan Neuropsychological Test Battery. Journal of the International Neuropsychological Society 5, 346–356, (1999).

24 Ritchie, K. et al. COGNITO: Computerised Assessment of Information Processing. J Psychol Psychother 4:2, 1000136, (2014).

25 Wechsler, D. WMS-IV Administration and Scoring Manual. (Pearson, 2010).

26 Wilson, B. A. et al. The Rivermead Behavioural Memory Test - Extended Version. (Thames Valley Test Company, 1999).

27 Wechsler, D. WAIS-IV Administration and Scoring Manual. (Pearson, 2010).

28 Nelson, H. E. & Willison, J. National Adult Reading Test (NART). (Nfer-Nelson Windsor, 1991).

29 Delis, D. C., Kaplan, E. & Kramer, J. H. Delis-Kaplan Executive Function System (D-KEFS) Examiner’s Manual. (The Psychological Corporation, 2001).

30 Roth, M., Huppert, F. A., Tym, E. & Mountjoy, C. Q. CAMDEX-R: The Cambridge Examination for Mental Disorders of the Elderly. (Cambridge University Press, 1998).

31 Cooper, C. Test the Nation: The IQ Book. (BBC Books, 2003).

32 Hampshire, A., MacDonald, A. & Owen, A. M. Hypoconnectivity and hyperfrontality in retired American football players. Sci Rep 3, 2972, (2013).

33 Wechsler, D. WAIS-IV Technical and Interpretive Manual. (Pearson, 2008).

